# Temporal trends of key preconception indicators among women in Northern Ireland, UK: An analysis of maternity healthcare data 2011-2021

**DOI:** 10.1101/2025.04.14.25325020

**Authors:** Emma H Cassinelli, Lisa Kent, Kelly-Ann Eastwood, Danielle AJM Schoenaker, Michelle C McKinley, Laura McGowan

**Affiliations:** Centre for Public Health, School of Medicine, Dentistry and Biomedical Sciences, Queen’s University Belfast, Belfast (United Kingdom); University Hospitals Bristol NHS Foundation Trust, Bristol (United Kingdom); School of Human Development and Health, Faculty of Medicine, University of Southampton, Southampton (United Kingdom); MRC Lifecourse Epidemiology Centre, University of Southampton, Southampton (United Kingdom); NIHR Southampton Biomedical Research Centre, University of Southampton and University Hospital Southampton NHS Foundation Trust, Southampton (United Kingdom)

**Keywords:** Preconception health and care, Health indicators, Epidemiology, Maternal health, Administrative data

## Abstract

**Background:** Optimising preconception health offers an opportunity to reverse unfavourable trends in modifiable risk factors and improve reproductive outcomes. This study aims to report the yearly prevalence of key biopsychosocial preconception indicators for over a decade, as reported at antenatal booking appointments in Northern Ireland (UK). The indicators include area-level deprivation, planned pregnancies, smoking, and body mass index (BMI) between 2011 and 2021, as well as pre- and early-pregnancy folic acid supplement use between 2015 and 2020.

**Methods:** This population-based study was conducted using annual routinely collected maternity data from the Northern Ireland Maternity System (NIMATS). R, accessed via the UK Secure eResearch Platform, was used to calculate yearly proportions. Multinomial regression models explored the relationship between each preconception indicator and year of booking appointment. Patient and Public Involvement and Engagement strategies were integrated throughout the study.

**Results:** The findings highlighted diverging trends over the study period; for example, smoking prevalence decreased (16.7%-9.6%), while obesity increased (e.g., obesity class I: 12.0%-16.1%). Preconception folic acid supplement use remained inadequate among women in NI, though the use of supplements containing 5mg of folic acid increased between 2015 and 2020 (400µg: 34.4%-30.03%; 5mg: 3.6%-5.0%).

**Conclusions:** Efforts are needed to reverse negative public health consequences of sub-optimal preconception health indicators. Notably, folic acid supplement use was predominantly initiated after conception, suggesting that a renewed focus is needed, particularly supporting women with the greatest need, such as those living in the most deprived areas.

## Introduction

Worldwide, population health has seen significant changes in recent decades, including increasing levels of obesity and associated co-morbidities such as diabetes.^1, 2, 3^ Behavioural factors influencing health such as insufficient physical activity have also been on the rise globally^4^, and evidence suggests that mental health disorders have increased globally since 1990.^5^ Optimising preconception health (i.e., the health of non-pregnant individuals of childbearing age) offers an opportunity to reverse unfavourable trajectories in modifiable risk factors and improve reproductive outcomes.^6^ Certain preconception indicators are more amenable to change than others, through individual behavioural modifications (e.g., smoking) or population-level initiatives (e.g., folic acid fortification).

In Northern Ireland (NI), one of four countries in the United Kingdom, data on key preconception indicators are recorded in the Northern Ireland Maternity System (NIMATS). It includes approximately 22,000 pregnancies each year from women cared for within the National Health Service (NHS) maternity services.^2^ Building on a previous study exploring the prevalence of key preconception indicators in NIMATS (2011-2021),^7^ an investigation of temporal trends was undertaken to gain further insight into preconception indicators and explore the implications for the delivery of preconception care. These insights can inform policymakers and healthcare professionals (HCPs) on the priority areas of need within the population, thereby assisting the development of future campaigns and interventions.^8^

### Aims

The current study aims to report the yearly prevalence of key preconception indicators, namely area-level deprivation, planned pregnancy, smoking, and body mass index (BMI), as recorded in NIMATS between 2011 and 2021 during antenatal booking appointments (i.e., the initial consultations pregnant women have with HCPs, which usually takes place by the 10^th^ or 12^th^ week of pregnancy).^9, 10^ The temporal trends of periconceptual folic acid supplement use were explored between 2015 and 2020, including a sub-analysis based on women’s area-level deprivation, age at booking, gravidity, planned pregnancy, and BMI. This sub-analysis focused on folic acid supplement use due to a higher-than-expected incidence of neural tube defects (NTDs) in the UK and Ireland compared to other countries in Europe since the early 1980s,^11, 12^ and due to recent UK Government legislation calls for mandatory folic acid fortification of non-wholemeal wheat flour.^13^ Additionally, there are ongoing debates regarding the optimal level of folic acid supplement use for the population and, therefore, a deeper understanding of this behaviour over time is pivotal. For example, while guidelines have so far recommended that women at high risk of an NTD, including women with obesity (BMI≥30kg/m^2^), should be advised to take 5mg of folic acid daily,^14^ recent suggestions have specified that women with a BMI≥25kg/m^2^ may not need the higher dosage of folic acid unless an additional co-morbidity is identified, for example diabetes or taking anti-epileptic medications.^15^ Because the investigations on folic acid supplement use trends have been limited in recent years and previously described as ‘patchy’ in the UK,^16, 17, 18^ this study aims to contribute to a better understanding of folic acid supplement use in NI, alongside other key biopsychosocial preconception indicators.

## Methods

A repeated cross-sectional population-based study was conducted using annual routinely collected maternity data from NIMATS. The selection of the indicators to analyse was informed by previous literature^19, 20^ and input from Patient and Public Involvement and Engagement (PPIE) representatives (previously reported).^7^ The analysed biopsychosocial indicators included planned pregnancy, smoking, and folic acid supplement use, representing behavioural factors, as well as BMI as a biological factor and deprivation quintiles as a social determinant of health. Further analyses exploring the temporal trends of periconceptual folic acid supplement use were conducted (i.e., based on women’s area-level deprivation, age at booking, gravidity, planned pregnancy, and BMI) due to concerns with the intake of this micronutrient in the UK. Other relevant indicators (i.e., alcohol consumption, diet quality, pre-existing physical and mental health conditions, and previous obstetric complications) were explored in the initial analyses, however these data are not reported due to significant data quality limitations identified.

Pregnancies with an antenatal booking appointment (and pregnancy outcome, such as a live birth) recorded between January 2011 and December 2021 were included. For the sub-analysis specifically investigating folic acid supplement use, pregnancies with a booking appointment between January 2015 and December 2020 were retained due to data availability. Details of the included indicators can be found in Supplementary material I.

### Analysis and statistical disclosure control

To avoid small counts (n<10), certain results were combined. For example, the ‘Missing’ values of folic acid supplement use were aggregated to ‘None’ and, instead of yearly prevalences, folic acid supplement use based on women’s BMI was presented as prevalences in 2015-2016, 2017-2018, and 2019-2021. Numerical restrictions were applied to the BMI variable, with values retained only if between 14-70 kg/m^2^, after exploring the pattern of individual data points. In the analyses on folic acid supplement use, only complete cases were retained due to disclosure controls.

After calculating the yearly prevalences of the included preconception indicators, multinomial logistic regression models explored the relationship between each indicator (outcome variables) and year of booking appointment (explanatory variable). Both unadjusted models and models adjusted for maternal age group and gravidity (i.e., the number of times a woman has been pregnant, including the current pregnancy) are presented. R,^21^ accessed via the UK Secure eResearch Platform, was used to conduct statistical analyses. Ethical approval and participant consent were not required.

### Patient and Public Involvement and Engagement

Members of the Healthy Reproductive Years advisory panel, which includes adults aged 18-45 years old residing in NI when recruited and belonging to different reproductive stages (e.g., planning for conception, recent parents), were involved throughout the study. Specifically, members were involved in the application for NIMATS data access (n=2) and prioritisation of the preconception indicators to analyse (n=11; previously reported).^7^ Further PPIE was conducted in collaboration with SureStart, a programme supporting parents with children under four years old living in disadvantaged areas in NI.

Women attended an in-person session to aid the interpretation of the findings relating specifically to folic acid supplement use, given the particular concerns with trends identified for this preconception indicator, and discussed ways to support and enable positive behaviour change before conception. The Guidance for Reporting Involvement of Patients and the Public (GRIPP) 2 checklist was used to report PPIE activities^22^ (Supplementary material II).

### Terminology

The term ‘woman’ is used to also include those who do not identify as women but have been or may become pregnant.^9^

## Results

The trends in key preconception indicators (2011-2021) were analysed across 255,117 pregnancies. Most pregnancies were from women living in the most deprived quintile (21.4%) and women who reported a planned pregnancy (70.6%). Less than half of the pregnancies were from women with a healthy BMI at booking (45.7%). The cohort used for the analyses on folic acid supplement use (2015-2020) included 132,205 pregnancies. Within this cohort, just under a third of pregnancies were primigravida (30.9%), and over a fifth were conceived by women living in the most deprived quintile (21.6%). Greater detail on women’s characteristics for the two cohorts is presented in Table 1. Regression models are presented in Tables 2a-2c (accompanying tables and figures in Supplementary material III, IV, and V).

**Table 1.**
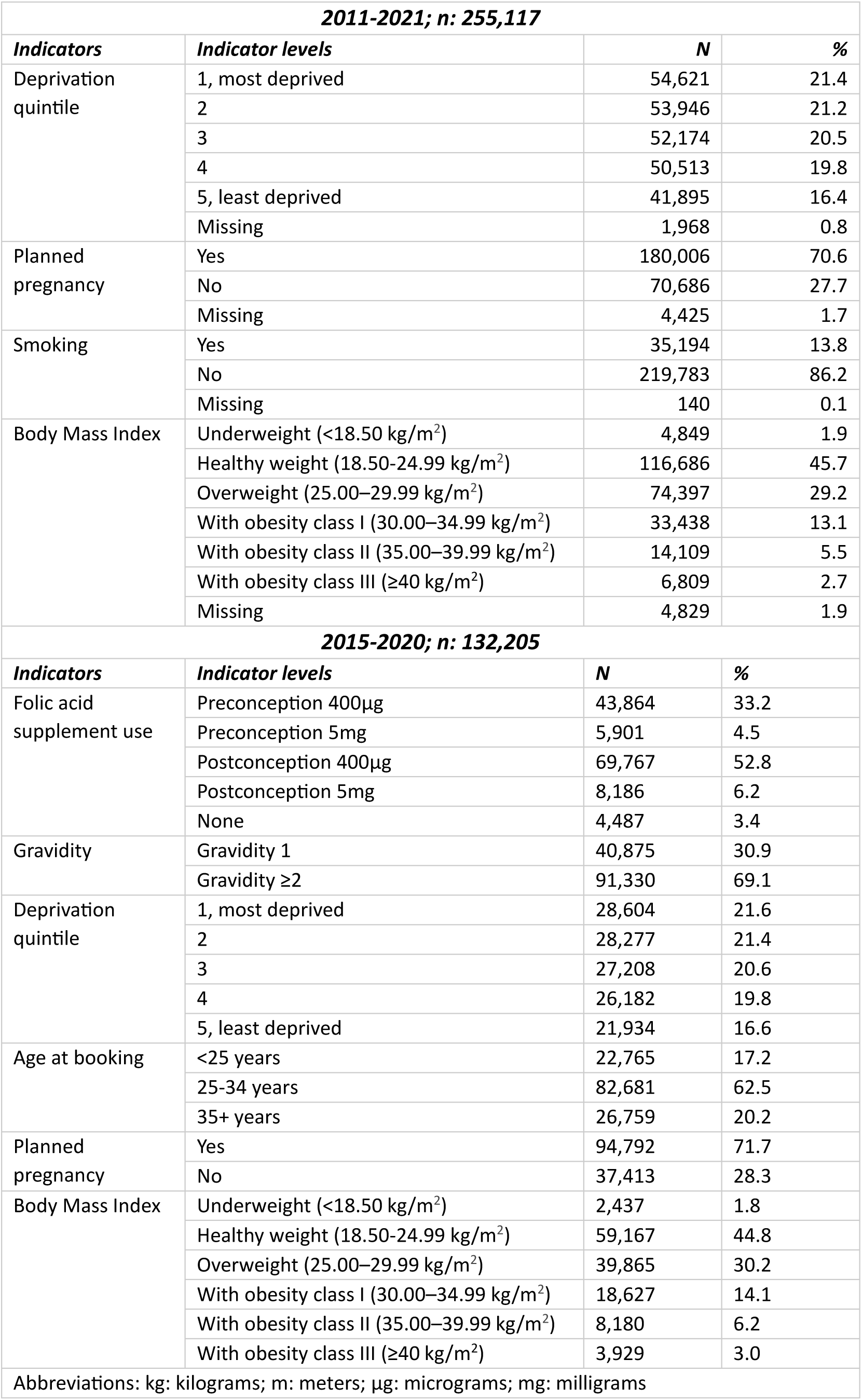
Characteristics of pregnancies in the 2011-2021 cohort (n: 255,117) and in the 2015-2020 cohort (n: 132,205).

**Table 2a.** Associations between year of booking interview and deprivation quintile, planned pregnancy, and smoking.

| *Year of booking* | *Deprivation quintile\** |  | *Planned pregnancy* |  | *Smoking* |  |
| --- | --- | --- | --- | --- | --- | --- |
|  | Unadjusted OR (95%CI) | Adjusted OR (95%CI) | Unadjusted OR (95%CI) | Adjusted OR (95%CI) | Unadjusted OR (95%CI) | Adjusted OR (95%CI) |
| 2011 | Ref | Ref | Ref | Ref | Ref | Ref |
| 2012 | 1.01 (0.98-1.04) | 0.99 (0.96-1.02) | **1.06 (1.02-1.10)** | 1.01 (0.97-1.06) | **0.99 (0.98-1.00)** | 0.96 (0.91-1.01) |
| 2013 | 1.01 (0.98-1.04) | 0.98 (0.95-1.02) | **1.08 (1.04-1.13)** | 1.01 (0.97-1.06) | **0.98 (0.98-0.99)** | **0.90 (0.86-0.95)** |
| 2014 | 0.99 (0.96-1.02) | **0.96 (0.93-0.99)** | 1.04 (1.00-1.08) | 0.95 (0.91-0.99) | **0.98 (0.97-0.98)** | **0.87 (0.83-0.92)** |
| 2015 | 1.01 (0.98-1.04) | 0.97 (0.94-1.00) | **1.08 (1.04-1.12)** | 0.97 (0.93-1.02) | **0.97 (0.96-0.97)** | **0.82 (0.78-0.86)** |
| 2016 | 1.02 (0.99-1.06) | 0.97 (0.94-1.00) | **1.08 (1.04-1.13)** | **0.95 (0.91-0.99)** | **0.96 (0.96-0.97)** | **0.81 (0.77-0.86)** |
| 2017 | 1.04 (1.00-1.07) | 0.98 (0.95-1.01) | 1.05 (1.01-1.09) | **0.90 (0.86-0.94)** | **0.96 (0.96-0.97)** | **0.82 (0.78-0.86)** |
| 2018 | 1.01 (0.98-1.04) | **0.94 (0.91-0.97)** | 1.02 (0.98-1.07) | **0.87 (0.83-0.91)** | **0.96 (0.95-0.97)** | **0.80 (0.76-0.84)** |
| 2019 | **1.05 (1.02-1.08)** | 0.97 (0.94-1.00) | 0.99 (0.95-1.03) | **0.82 (0.79-0.86)** | **0.94 (0.94-0.95)** | **0.69 (0.65-0.72)** |
| 2020 | 1.03 (1.00-1.06) | **0.95 (0.92-0.98)** | **1.06 (1.01-1.10)** | **0.86 (0.82-0.90)** | **0.92 (0.91-0.93)** | **0.52 (0.49-0.55)** |
| 2021 | **1.05 (1.01-1.10)** | 0.96 (0.93-1.00) | **1.11 (1.06-1.17)** | **0.90 (0.85-0.95)** | **0.91 (0.90-0.92)** | **0.450.42-0.49)** |

\*Range from 1, most deprived, to 5, least deprived. Abbreviations: OR: Odds ratio; CI: Confidence Interval. Adjusted: deprivation quintile + age group + gravidity. Results in bold represent $p < 0.01$ .

**Table 2b.** Associations between year of booking interview and BMI categories.

| *Year of booking* | *BMI categories* |  |  |  |  |  |  |  |  |  |  |  |
| --- | --- | --- | --- | --- | --- | --- | --- | --- | --- | --- | --- | --- |
|  | Unadjusted OR (95%CI) |  |  |  |  |  | Adjusted OR (95%CI) |  |  |  |  |  |
|  | Underweight | Healthy weight | Overweight | With obesity I | With obesity II | With obesity III | Underweight | Healthy weight | Overweight | With obesity I | With obesity II | With obesity III |
| 2011 | Ref | Ref | Ref | Ref | Ref | Ref | Ref | Ref | Ref | Ref | Ref | Ref |
| 2012 | 1.12 (0.99-1.27) | Ref | 1.02 (0.98-1.06) | 1.02 (0.97-1.08) | 1.03 (0.95-1.13) | 1.12 (0.99-1.27) | 1.16 (1.02-1.31) | Ref | 1.02 (0.97-1.06) | 1.02 (0.96-1.08) | 1.03 (0.95-1.13) | 1.12 (0.99-1.27) |
| 2013 | 1.04 (0.91-1.18) | Ref | 1.04 (0.99-1.08) | 1.03 (0.98-1.09) | 1.07 (0.98-1.17) | **1.11 (0.98-1.25)** | 1.09 (0.95-1.23) | Ref | 1.03 (0.99-1.07) | 1.03 (0.97-1.09) | 1.06 (0.98-1.16) | 1.10 (0.97-1.24) |
| 2014 | 1.01 (0.89-1.15) | Ref | 1.03 (0.99-1.07) | 1.07 (1.01-1.13) | **1.17 (1.07-1.27)** | **1.22 (1.08-1.38)** | 1.07 (0.94-1.22) | Ref | 1.02 (0.98-1.06) | 1.06 (1.00-1.13) | **1.16 (1.07-1.26)** | **1.21 (1.07-1.36)** |
| 2015 | 1.12 (0.98-1.27) | Ref | **1.10 (1.06-1.15)** | **1.12 (1.05-1.18)** | **1.21 (1.12-1.32)** | **1.26 (1.12-1.43)** | **1.20 (1.06-1.36)** | Ref | **1.09 (1.04-1.13)** | **1.11 (1.05-1.17)** | **1.21 (1.11-1.31)** | **1.25 (1.11-1.41)** |
| 2016 | 1.01 (0.89-1.15) | Ref | **1.13 (1.08-1.18)** | **1.23 (1.16-1.30)** | **1.38 (1.27-1.50)** | **1.49 (1.32-1.67)** | 1.10 (0.97-1.26) | Ref | **1.11 (1.07-1.16)** | **1.22 (1.15-1.29)** | **1.37 (1.26-1.49)** | **1.47 (1.30-1.65)** |
| 2017 | 1.07 (0.94-1.22) | Ref | **1.13 (1.09-1.18)** | **1.26 (1.19-1.33)** | **1.46 (1.34-1.58)** | **1.41 (1.25-1.59)** | **1.18 (1.03-1.35)** | Ref | **1.12 (1.07-1.17)** | **1.25 (1.18-1.32)** | **1.45 (1.33-1.57)** | **1.39(1.23-1.57)** |
| 2018 | 0.93 (0.81-1.07) | Ref | **1.15 (1.10-1.20)** | **1.35 (1.27-1.43)** | **1.54 (1.42-1.68)** | **1.61 (1.43-1.82)** | 1.04 (0.90-1.19) | Ref | **1.13 (1.09-1.18)** | **1.34 (1.27-1.42)** | **1.54 (1.41-1.67)** | **1.60 (1.42-1.80)** |
| 2019 | 1.03 (0.90-1.18) | Ref | **1.25 (1.19-1.30)** | **1.49 (1.41-1.58)** | **1.63 (1.50-1.77)** | **1.81 (1.61-2.04)** | 1.16 (1.01-1.33) | Ref | **1.23 (1.18-1.28)** | **1.49 (1.41-1.58)** | **1.63 (1.50-1.78)** | **1.80 (1.60-2.03)** |
| 2020 | 0.96 (0.83-1.10) | Ref | **1.28 (1.23-1.34)** | **1.50 (1.42-1.59)** | **1.91 (1.76-2.07)** | **2.07 (1.84-2.32)** | 1.10 (0.95-1.27) | Ref | **1.26 (1.21-1.32)** | **1.50 (1.41-1.58)** | **1.90 (1.75-2.06)** | **2.04 (1.82-2.29)** |
| 2021 | 0.94 (0.78-1.12) | Ref | **1.35 (1.28-1.43)** | **1.69 (1.58-1.81)** | **2.02 (1.83-2.22)** | **2.47 (2.16-2.82)** | 1.09 (0.91-1.30) | Ref | **1.32 (1.25-1.39)** | **1.68 (1.56-1.80)** | **2.00 (1.82-2.21)** | **2.42 (2.12-2.77)** |
| Abbreviations: OR: Odds ratio; CI: Confidence Interval. Adjusted: deprivation quintile + age group + gravidity. Results in bold represent $p<0.01$. | | | | | | | | | | | | |

**Table 2c.** Associations between year of booking interview and folic supplement use.

| Year of booking | Folic acid supplement use (timing) |  |  |  |  |  | Folic acid supplement use (dosage) |  |  |  |  |  |
| --- | --- | --- | --- | --- | --- | --- | --- | --- | --- | --- | --- | --- |
|  | Unadjusted OR (95%CI) |  |  | Adjusted OR (95%CI) |  |  | Unadjusted OR (95%CI) |  |  | Adjusted OR (95%CI) |  |  |
|  | Pre | Post | None | Pre | Post | None | 400 µg | 5 mg | None | 400 µg | 5 mg | None |
| 2015 | Ref | Ref | Ref | Ref | Ref | Ref | Ref | Ref | Ref | Ref | Ref | Ref |
| 2016 | Ref | 0.99 (0.95-1.02) | 0.90 (0.82-1.00) | Ref | 1.00 (0.96-1.04) | 0.93 (0.84-1.03) | Ref | **1.19 (1.11-1.27)** | 0.93 (0.84-1.02) | Ref | **1.19 (1.11-1.27)** | 0.94 (0.85-1.04) |
| 2017 | Ref | 1.01 (0.97-1.05) | **0.86 (0.77-0.95)** | Ref | 1.03 (0.99-1.07) | 0.89 (0.80-0.99) | Ref | **1.48 (1.39-1.58)** | 0.88 (0.80-0.98) | Ref | **1.47 (1.38-1.57)** | 0.90 (0.82-1.00) |
| 2018 | Ref | 0.99 (0.95-1.03) | 0.91 (0.82-1.01) | Ref | 1.01 (0.98-1.06) | 0.96 (0.86-1.06) | Ref | **1.55 (1.45-1.65)** | 0.96 (0.87-1.06) | Ref | **1.54 (1.44-1.64)** | 0.99 (0.89-1.09) |
| 2019 | Ref | 1.01 (0.97-1.05) | 0.88 (0.80-0.98) | Ref | **1.06 (1.01-1.10)** | 0.95 (0.86-1.06) | Ref | **1.82 (1.71-1.94)** | 0.93 (0.84-1.03) | Ref | **1.81 (1.70-1.92)** | 0.98 (0.88-1.08) |
| 2020 | Ref | **1.15 (1.10-1.19)** | 0.99 (0.89-1.10) | Ref | **1.21 (1.16-1.26)** | **1.09 (0.98-1.21)** | Ref | **1.76 (1.65-1.88)** | 0.96 (0.87-1.07) | Ref | **1.74 (1.64-1.86)** | 1.02 (0.92-1.12) |
| Abbreviations: Odds ratio; CI: Confidence Interval; Pre: Preconception; Post: Postconception; µg: micrograms; mg: milligrams. Adjusted: deprivation quintile + age group + gravidity. Results in bold represent $p < 0.01$. | | | | | | | | | | | | |

Fewer pregnancies were recorded in the least deprived quintile compared to the most deprived quintile. No notable variations in deprivation were observed between 2011 and 2021 (q1: 21.8%-21.4%; q5: 16.1%-17.4%), with the regression model adjusted for maternal age and gravidity not reaching statistical significance in most years. The yearly prevalence of pregnancies reported as planned slightly increased between 2011 and 2021 (70.8%-73.0%), though an adjusted OR (aOR) of 0.90 (95%CI 0.85-0.95, *p<*0.01) was observed. Rates of smoking declined from 16.7% to 9.6%; women with a booking in 2021 had significantly lower odds of reporting smoking than those with a booking in 2011 (aOR 0.45, 95%CI 0.42-0.49, *p<*0.01). A decline was observed in pregnancies conceived by women with a healthy BMI (50.3%-39.9%), with opposing trends (i.e., increases) shown for women with a BMI≥25kg/m^2^ (overweight: 29.0%-31.0%; obesity class I: 12.0%-16.1%; obesity class II: 4.6%-7.4%; obesity class III: 2.1%-4.1%). These results were confirmed in the adjusted regression models; for example, women with a booking in 2021 were more likely to have class III obesity than women with a booking in 2011 (aOR 2.42, 95% CI 2.12-2.77, *p<*0.01).

Analyses suggested an overall decrease in the preconception use of supplements containing 400μg of folic acid between 2015 and 2020 (34.4%-30.0%) and a slight increase in preconception use of supplements containing 5mg (3.6%-5.0%). Postconception use of 400μg supplements declined between 2015 and 2019 (54.3%-51.4%) then slightly increased to 53.94% in 2020. Postconception use of 5mg supplements steadily increased (4.0%-7.7%). Women with a booking in 2020 were more likely to report postconception supplement use rather than preconception (aOR 1.21, 95%CI 1.16-1.26, *p<*0.01) and report using supplements with 5mg rather than 400μg (aOR 1.74, 95%CI 1.64-1.86, *p<*0.01), compared to women with a booking in 2015.

Preconception use of 400μg folic acid supplements declined between 2015 and 2020 (34.4%-30.0%), with the largest percentage difference per subgroup observed among women from the most deprived quintile (24.3%-19.5%), women aged 35+ years (44.0%-37.2%), subsequent pregnancies (34.3%-28.9%), planned pregnancies (45.4%-39.5%), and women with obesity class III (20.0%-10.9%).

Postconception folic acid supplement use of 400μg exhibited no major variations across deprivation quintiles, though it was highest in women from the most deprived quintile. Based on maternal age, it was highest in women younger than 25 years, but in the last year (2019-2020) women aged 25–34 years exhibited the greatest increase (49.3%-52.8%). In first-time pregnancies, it was varied across the studied years and lowest in 2016 (51.9%) and 2019 (51.7%); in subsequent pregnancies, a decline was observed between 2015 and 2019 (53.8%-51.2%), followed by a slight increase between 2019 and 2020 (51.2%-53.9%). Postconception use of 400μg folic acid supplements decreased among unplanned pregnancies between 2015 and 2020 (79.2%-75.5%). In pregnancies conceived by women with underweight, healthy weight, and overweight, it slightly increased throughout the study period (62.5%-64.4%, 54.1%-54.9%, and 54.7%-55.6%, respectively).

The use of supplements containing 5mg of folic acid before or after conception increased between 2015 and 2020, with no notable differences across deprivation quintiles. It increased for all age groups, though in the last year preconception supplement use of 5mg declined among women aged 35+ years (9.0%-7.1%). The use of 5mg supplements also increased irrespective of gravidity. Regarding pregnancy planning status, the largest percentage change was observed among unplanned pregnancies, where postconception use of 5mg supplements increased (6.0%-10.2%). The use of supplements with 5mg of folic acid steadily increased among pregnancies by women with obesity before (e.g., class III: 8.3%-15.7%) and after conception (e.g., class III: 23.1%-38.0%). Figures illustrating these trends in folic acid supplement use are included in Supplementary material VI.

### Patient and Public Involvement and Engagement

During a drop-in breastfeeding support group session organised by SureStart, nine women informed the interpretation of the present findings reporting periconceptual folic acid supplement use and participated in related discussions. Some women acknowledged the importance of folic acid supplement use, but often lacked awareness of dosage options and benefits. For advice, they discussed relying on HCPs, family, and online platforms, though trust in social media advice varied. Suggested improvements included greater GP involvement, personalised care, and educational initiatives (details in Supplementary material VII).

## Discussion

The present study explored temporal trends in key preconception indicators in NI. Many changes were observed in the prevalence of indicators between 2011 and 2021, with trends suggesting both positive changes (e.g., reductions in reported smoking) and areas of public health concern (e.g., increasing obesity rates). Certain changes were negligible, such as those observed among planned pregnancies. Investigations of folic acid supplement use between 2015 and 2020 highlighted groups of women in greatest need of support, including women under 25 years of age, women living in the most deprived quintiles, and women who did not report having a planned pregnancy.

### Trends in key preconception indicators

Trends in area-level deprivation remained relatively constant between 2011 and 2021, potentially indicating limited positive changes in tackling inequalities, however the proportion of planned pregnancies slightly increased. There may be many reasons behind this trend, including delayed maternal age for childbearing^23^ and the availability of contraception,^2^ though these trends may warrant further research (e.g., investigating the trends in planned pregnancies across subsequent pregnancies conceived by the same woman). Given the retrospective nature of the question used to collect data on pregnancy planning in NIMATS, social desirability and recall bias should also be considered as a possible influencing factor. Promisingly, results indicated decreasing rates of reported smoking. These trends, which echo existing evidence from NI,^2^ may be influenced by adults’ high level of perceived importance placed on preconception smoking cessation and the significant public health investment into smoking cessation services.^24^ However, though not measured in the present analyses, the proportion of women of reproductive age reporting using e-cigarettes has been rising in NI.^25^ Of public health concern and echoing previous evidence,^1, 2^ this study showed a decreasing prevalence of pregnancies conceived by women with a healthy BMI. The associated increasing prevalence of pregnancies by women with obesity is in line with the rising trends in the number of adults living with obesity in NI^25^ and globally.^3^ Meaningful efforts are needed to address these trends given the evidence that living with obesity can impact fertility, time to conceive, and increase the risk of adverse maternal and infant outcomes.^26^

Emerging concerns about periconceptual folic acid supplement use and its known link to NTDs^14^ prompted a more in-depth investigation of this indicator using available data over a 5-year period (2015-2020). Notably, preconception supplement use of 400μg of folic acid declined. Specifically, though it was lowest in the most deprived quintile, it declined across all area-level deprivation quintiles. Echoing previous literature,^27, 28, 29^ it was also low in pregnancies conceived by women aged <25 years and unplanned pregnancies. A pronounced decline in preconception use of 400μg folic acid supplements was observed among pregnancies by women with obesity, especially class III. This may reflect a positive shift, as women with obesity are typically prescribed the higher dose of folic acid (5mg/day),^14^ which increased over the study period. Postconception use of 400μg also declined until 2019, though trends exhibited a slight increase in 2020. Use was high among pregnancies conceived by women living in the most deprived quintile, with minimal variation over time, women aged <25 years, and unplanned pregnancies. A trend demonstrating a slightly increasing postconception use of 400μg supplements was noted in recent years among older women and planned pregnancies. The observed sub-optimal supplement use in preparation for subsequent pregnancies, as previously reported,^29^ indicates the need for improved interconception care in NI.

Preconception and postconception use of supplements with 5mg of folic acid increased over the timeperiod regardless of deprivation quintile, age, gravidity, and planned pregnancy, although it was often more pronounced in the postconception period. This trend was particularly evident among women with obesity, potentially due to the rising prevalence of overall obesity. Women with obesity, being at an increased risk of co-morbidities such as diabetes and hypertension,^26^ may have more opportunities to interact with HCPs, potentially facilitating the prescription of high-dose supplements. However, this cannot be concluded from the current study. In NI, the Weigh to a Healthy Pregnancy programme, which aims to support women with a BMI>38kg/m^2^ in managing their weight during pregnancy,^30^ may also contribute to information delivery regarding folic acid supplement use, particularly in subsequent pregnancies.

The current practice in the UK of recommending different doses of folic acid based on women’s risk of NTDs has been debated. The availability of supplements containing 5mg of folic acid only with a medical prescription has been contested, resulting in suggestions of standardising the daily recommended dosage to 4mg or 5mg without requiring a prescription.^31^ Moreover, while the analyses were conducted when guidelines recommended that women with obesity should take 5mg of folic acid,^14^ this too is under consideration. For example, the 2025 National Institute for Health and Care Excellence (NICE) guideline on maternal and child nutrition^15^ specified that women with a BMI≥25kg/m^2^ planning to conceive or in the first 12 weeks of pregnancy do not need to take supplements containing more than 400μg of folic acid, unless they have other specific health factors (e.g., having diabetes, taking anti-epileptic medications, having had a previous pregnancy complicated by an NTD). However, given that there is also evidence suggesting that women with overweight or obesity may be at a higher risk of having NTD-affected pregnancies, the Royal College of Obstetricians and Gynaecologists currently advises women with obesity wishing to become pregnant to take 5mg folic acid supplements.^32^ Overall, these differences highlight the continuing debate over optimal supplementation.

### Implications and future research

Examining national maternity data with significant population coverage has highlighted key areas of concern for population health, such as a need for addressing increasing rates of maternal BMI. Although beyond the scope of these analyses, population health data indicate increasing BMI trends also in males.^33^ This suggests that additional research is warranted to explore the implications of both maternal and paternal obesity on preconception health, especially given that evidence indicates increased risks.^34^

Within this study, a need for supporting appropriate preconception folic acid supplement use was also observed. It should be noted that women’s reporting of supplement use does not ensure their adherence to recommendations (e.g., daily intake for at least 12 weeks before and after conception),^14^ and that among those reporting postconception use, the timing of initiation after conception could not be determined (e.g., shortly after conception or just before the booking appointment). Therefore, future studies should strive to improve the recording of self-reported health behaviours such as supplement use, reporting timeframes (e.g., the weeks or months before conception when use was initiated), and adherence (e.g., daily intake).^35^ Furthermore, future assessments of supplement use could provide a better understanding without relying on self-reported measures (e.g., prescription records, biomarker assessments). Particularly vulnerable groups such as young women and women from areas of greatest deprivation may benefit from targeted interventions, including peer-delivered interventions^17^ and provision of free supplements,^18^ though population-level approaches are also recommended. Reservations about the effectiveness and long-term impacts of campaigns have been expressed in the literature, as well as concerns about interventions’ potential to exacerbate health inequalities.^18^ Healthcare professionals such as GPs, gynaecologists, and obstetricians may be especially well positioned for the provision of advice and recommend uptake^18, 27^, but the public’s lack of knowledge of preconception care and infrequent contact with HCPs before conceiving may be barriers to consider, as well as lack of time and resources for HCPs.^36, 37^ Alongside individualised HCP support, emerging technologies and social media may be harnessed to further promote preconception health, alongside input from PPIE representatives.

### Patient and Public Involvement and Engagement

The PPIE activities were overall well received by public members. Topics discussed by women from the SureStart programme included the reluctance to openly talk about pregnancy intentions before conception, retrieval of advice online or from HCPs, and desire to receive reliable and personalised advice.^37^ These discussions involved women who already had children and were breastfeeding, but other groups of women may report different views or have a different knowledge base of preconception health. Future research and initiatives should be informed by the views expressed by women with varied lived experiences of the preconception period.

### Strengths and limitations

This study presents novel findings from pregnancy-level analyses using routinely-collected maternity data across NI over an 11-year period to enhance the current epidemiological understanding of preconception health.

Some variables used in the analyses were self-reported, thereby potentially introducing recall and social desirability bias. Ethnicity is a factor that has been shown to be associated with preconception health,^18, 29^ though due to the limited ethnic diversity in NI^38^ and disclosure controls, it was not included in the present analyses. Similarly, factors including educational attainment were not directly explored in the present study due to data unavailability. However, the calculation of the deprivation quintiles used in this study, based on the NI Multiple Deprivation Measure (NIMDM) 2017, also accounts for women’s employment, education, skills, and training.^39^ Other factors that have been associated with preconception health, such as locality of residence, housing, immigrant status, and access to healthcare services^29, 35^ may warrant further investigations. The COVID-19 pandemic may also have affected childbearing behaviours, for example by affecting mental health and distress,^40^ or access to folic acid supplements. Because the data included in these analyses covered pregnancies with a booking appointment until 2020 or 2021, there were insufficient data collected following the outbreak to draw firm conclusions on its impact.

## Conclusions

This study offers an overview of key preconception indicators over time in NI, as recorded in routinely-collected maternity data. Key findings reveal both improving and concerning trends: while smoking rates declined, obesity increased. Additionally, analyses showed that preconception folic acid supplement use is still inadequate among women in NI, though the use of 5mg supplements increased, trend potentially associated with the increasing prevalence of women with obesity. A renewed focus on particular subgroups of women, including young women and women living in the most deprived areas, is recommended as a result of this study’s findings.

## Funding

This work is supported by Queen’s University Belfast as part of a PhD studentship for EHC. EHC is funded by the Department for the Economy NI. The funder has no specific role in the conceptualisation, design, data collection, analysis, decision to publish, or preparation of the manuscript. DAJMS is supported by the National Institute for Health and Care Research (NIHR) through an NIHR Advanced Fellowship (NIHR302955) and the NIHR Southampton Biomedical Research Centre (NIHR203319). Disclaimer: The views expressed are those of the author(s) and not necessarily those of the NIHR or the Department of Health and Social Care.

## Conflicts of Interests

No interests to disclose.

## Data Availability

The datasets used in the analyses in the present study can be accessed via the Honest Broker Service secure data environment following a successful application.

## Acknowledgements

The authors would like to acknowledge the help provided by the staff of the Honest Broker Service (HBS) within the Business Services Organisation Northern Ireland (BSO). The HBS is funded by the BSO and the Department of Health (DoH). The authors alone are responsible for the interpretation of the data and any views or opinions presented are solely those of the author and do not necessarily represent those of the BSO. The authors would like to acknowledge the contribution of the Patient and Public Involvement and Engagement ‘Healthy Reproductive Years’ panel for their help and support.

## Key points

- Temporal trends of key biopsychosocial preconception indicators among women in Northern Ireland (UK) are presented, between 2011 and 2021 or between 2015 and 2020, based on data availability in the Northern Ireland Maternity System dataset.
- Both improving and concerning trends were identified; for example, while smoking rates declined, obesity increased during the study period.
- Periconceptual folic acid supplement use differed across the population. For example, women living in the most deprived quintile and younger women were more likely to report reliance on postconception, rather than preconception, folic acid supplement use (as per recommendations), compared to women living in the least deprived quintile or women aged 25 years or over, respectively.
- The discussion of findings with Patient and Public Involvement and Engagement representatives identified a level of confusion on certain aspects of folic acid supplement use recommendations, reluctance to openly talk about pregnancy intentions before conception, and desire to receive personalised preconception advice.

## Notes

### Competing Interest Statement

The authors have declared no competing interest.

### Funding Statement

This work was supported by Queens University Belfast as part of a PhD studentship for EHC. EHC was funded by the Department for the Economy NI. The funder has no specific role in the conceptualisation, design, data collection, analysis, decision to publish, or preparation of the manuscript. DS is supported by the National Institute for Health and Care Research (NIHR) through an NIHR Advanced Fellowship (NIHR302955) and the NIHR Southampton Biomedical Research Centre (NIHR203319). Disclaimer: The views expressed are those of the author(s) and not necessarily those of the NIHR or the Department of Health and Social Care.

### Author Declarations

Applications to the Honest Broker Service are considered by the Health and Social Care (HSC) Data Access Committee. No additional approvals, such as by HSC Research Ethics Committee, were required for the project. Researchers working on the project signed the mandatory Research Data Access Agreement, and successfully completed the Office for National Statistics Safe Researcher training.

