## Supplementary Materials for "Temporal trends of key preconception indicators among women in Northern Ireland, UK: An analysis of maternity healthcare data 2011-2021"

### Supplementary material I

| *Indicator* | *How is it recorded in NIMATS?* | *How is it presented in this study?* |
|---|---|---|
| Deprivation quintile* | Area-level deprivation was estimated using NIMDM 2017, which aggregates the ranking of seven specific domains into a single ranking. These weighted domains are: 25% income, 25% employment, 15% health and disability, 15% education, skills and training, 10% access to services, 5% living environment, and 5% crime and disorder[1]. Maternal data in NIMATS were linked to the NIMDM by the super output area code[2]. | Area-level deprivation was categorised into quintiles, where the first quintile represented the most deprived areas and the fifth the least deprived ones. |
| Planned pregnancy | Planned pregnancies were recorded as a binary Yes/No measure at booking. | Planned pregnancies were presented as a Yes/No measure. |
| Smoking | Smoking status was calculated based on the reported number of cigarettes smoked per day at booking. | Smoking status was presented as a Yes/No measure. |
| BMI | BMI was derived by clinically measured weight and height recorded at booking. | Women were divided into: underweight (<18.49 kg/m$^2$), healthy weight (18.50–24.99 kg/m$^2$), overweight (25.00–29.99 kg/m$^2$), with obesity class I (30.00–34.99 kg/m$^2$), with obesity class II (35.00–39.99 kg/m$^2$), and with obesity class III (≥40.00 kg/m$^2$)[3]. Recordings of BMI were included if in the range of 14-70 kg/m$^2$. |
| Folic acid supplement use | Folic acid supplement use was categorised based on both the timing (i.e., preconception and postconception) and dose reported (i.e., 400µg or 5mg). The data included for this measure were only pertaining to pregnancies with a booking appointment date after 01/12/2014, because of changes in the recording in NIMATS. | Folic acid supplement use was presented as Preconception 400µg, Preconception 5mg, Postconception 400µg, Postconception 5mg, and None. |

*Not recorded in NIMATS; postcodes were used to discern deprivation quintiles.
Abbreviations: BMI: Body Mass Index; kg: kilogram; m: metre; mg: milligram; NIMATS: Northern Ireland Maternity System; NIMDM: Northern Ireland Multiple Deprivation Measure; µg: microgram.

### Supplementary Material II

Table 1. Guidance for Reporting Involvement of Patients and the Public (GRIPP) short-form checklist.

| ***Section and topic*** | ***Item*** | ***Reported on page No*** |
|---|---|---|
| 1: Aim | Report the aim of PPIE in the study | 5-6 |
| 2: Methods | Provide a clear description of the methods used for PPIE in the study | 5-6 |
| 3. Study results | Outcomes—Report the results of PPIE in the study, including both positive and negative outcomes | 8-9 (and Supplementary material VII) |
| 4. Discussion and conclusion | Outcomes—Comment on the extent to which PPIE influenced the study overall. Describe positive and negative effects | 13 |
| 5. Reflections/critical perspective | Comment critically on the study, reflecting on the things that went well and those that did not, so others can learn from this experience | 13 |
| Abbreviation: PPI Patient and Public Involvement and Engagement. | | |

### Supplementary material III

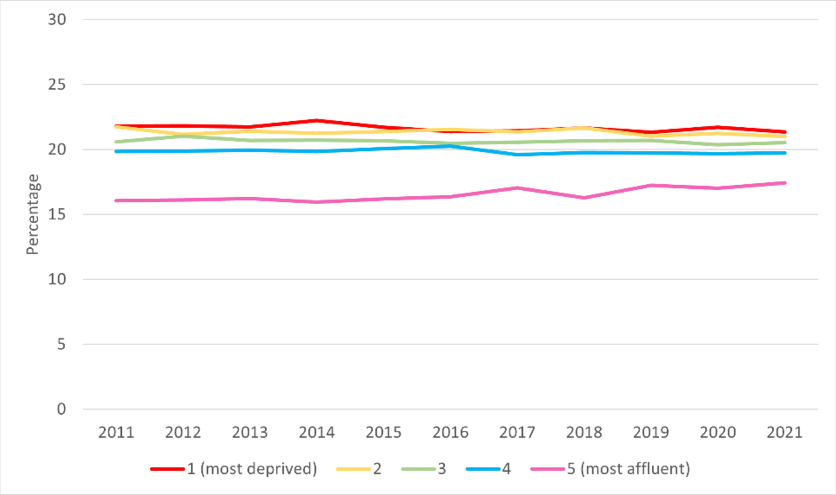


Figure 1a. Trends of deprivation quintiles.

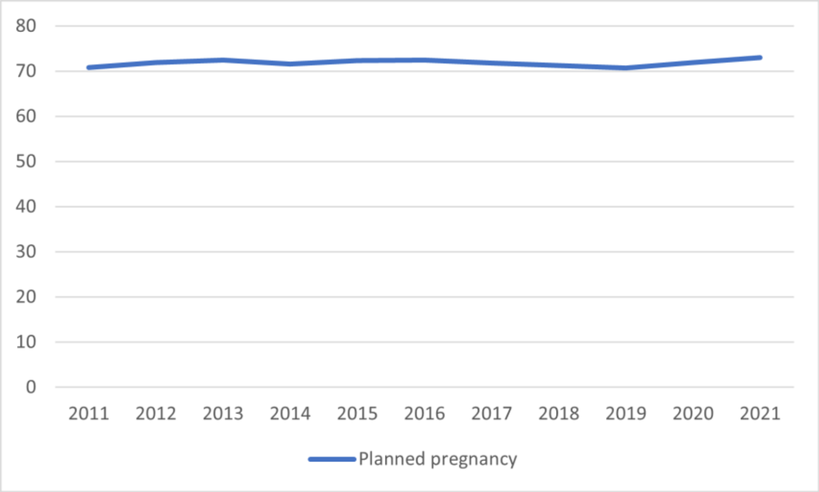


Figure 1b. Trends of reported planned pregnancies.

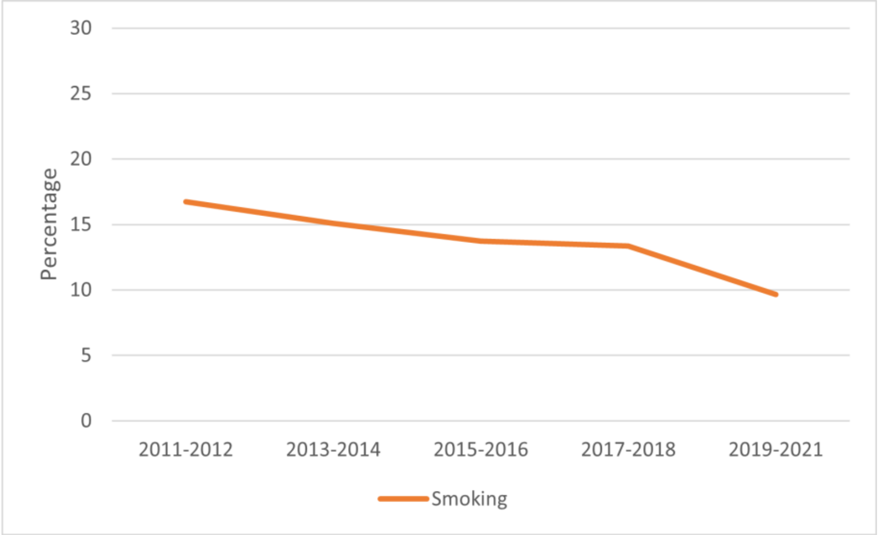


Figure 1c. Trends of reported smoking.

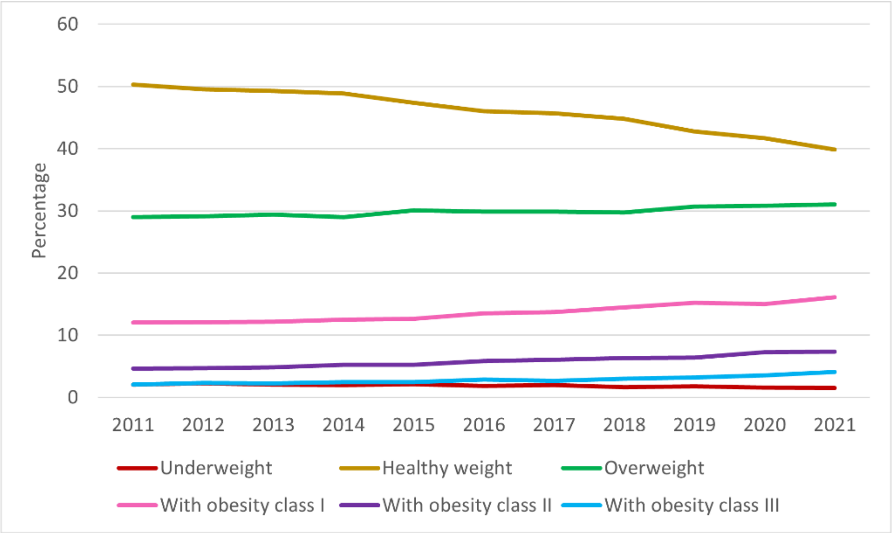


Figure 1d. Trends of BMI categories.

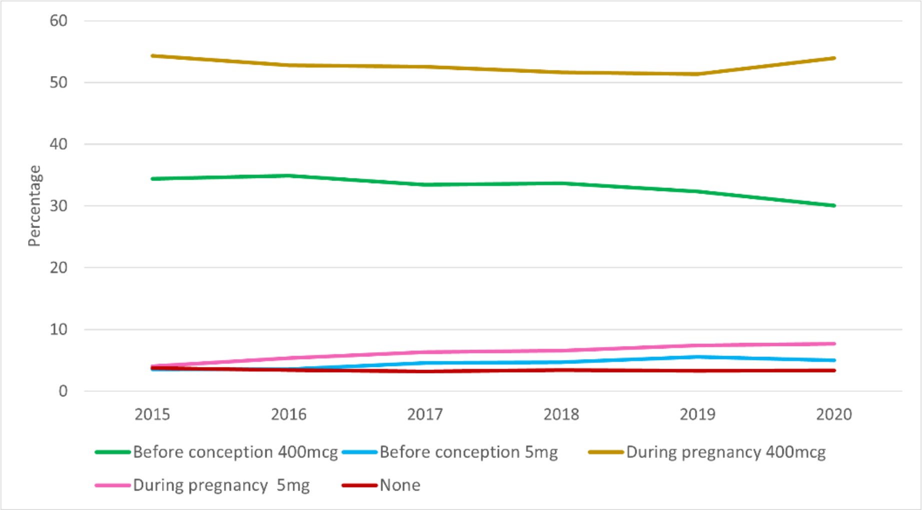


Figure 1e. Trends of reported periconception folic acid supplement use.

### Supplementary material IV

Table 1. Trends in deprivation (years: 2011-2021; data source: NIMATS).

| *Year of booking* | *Indicator level* | *N* | *%* |
|---|---|---|---|
| 2011 | 1 | 5367 | 21.79 |
| 2012 | | 5250 | 21.82 |
| 2013 | | 5190 | 21.73 |
| 2014 | | 5290 | 22.24 |
| 2015 | | 5174 | 21.71 |
| 2016 | | 4979 | 21.38 |
| 2017 | | 4826 | 21.44 |
| 2018 | | 4778 | 21.66 |
| 2019 | | 4621 | 21.31 |
| 2020 | | 4630 | 21.7 |
| 2021 | | 2320 | 21.35 |
| 2011 | 2 | 5355 | 21.74 |
| 2012 | | 5095 | 21.17 |
| 2013 | | 5113 | 21.41 |
| 2014 | | 5052 | 21.24 |
| 2015 | | 5094 | 21.37 |
| 2016 | | 5016 | 21.54 |
| 2017 | | 4809 | 21.36 |
| 2018 | | 4776 | 21.65 |
| 2019 | | 4561 | 21.03 |
| 2020 | | 4531 | 21.24 |
| 2021 | | 2280 | 20.99 |
| 2011 | 3 | 5068 | 20.58 |
| 2012 | | 5061 | 21.03 |
| 2013 | | 4940 | 20.68 |
| 2014 | | 4930 | 20.73 |
| 2015 | | 4924 | 20.66 |
| 2016 | | 4768 | 20.48 |
| 2017 | | 4630 | 20.56 |
| 2018 | | 4555 | 20.65 |
| 2019 | | 4488 | 20.69 |
| 2020 | | 4346 | 20.37 |
| 2021 | | 2230 | 20.53 |
| 2011 | 4 | 4887 | 19.84 |
| 2012 | | 4783 | 19.88 |
| 2013 | | 4767 | 19.96 |
| 2014 | | 4720 | 19.85 |
| 2015 | | 4781 | 20.06 |
| 2016 | | 4715 | 20.25 |
| 2017 | | 4413 | 19.60 |
| 2018 | | 4359 | 19.76 |
| 2019 | | 4280 | 19.73 |

| | | | |
|---|---|---|---|
| 2020 | | 4196 | 19.67 |
| 2021 | | 2142 | 19.72 |
| 2011 | 5 | 3953 | 16.05 |
| 2012 | | 3876 | 16.11 |
| 2013 | | 3875 | 16.22 |
| 2014 | | 3792 | 15.94 |
| 2015 | | 3860 | 16.20 |
| 2016 | | 3807 | 16.35 |
| 2017 | | 3836 | 17.04 |
| 2018 | | 3590 | 16.28 |
| 2019 | | 3738 | 17.24 |
| 2020 | | 3630 | 17.02 |
| 2021 | | 1892 | 17.42 |

Table 2. Trends in planned pregnancy (years: 2011-2021; data source: NIMATS).

| ***Year of booking*** | ***Indicator level*** | ***N*** | ***%*** |
|---|---|---|---|
| 2011 | Yes | 17173 | 70.81 |
| 2012 | | 17036 | 71.94 |
| 2013 | | 17106 | 72.44 |
| 2014 | | 16826 | 71.59 |
| 2015 | | 17030 | 72.36 |
| 2016 | | 16635 | 72.44 |
| 2017 | | 16034 | 71.75 |
| 2018 | | 15714 | 71.28 |
| 2019 | | 15295 | 70.67 |
| 2020 | | 15308 | 71.91 |
| 2021 | | 7914 | 72.98 |
| 2011 | No | 7079 | 29.19 |
| 2012 | | 6645 | 28.06 |
| 2013 | | 6509 | 27.56 |
| 2014 | | 6676 | 28.41 |
| 2015 | | 6504 | 27.64 |
| 2016 | | 6328 | 27.56 |
| 2017 | | 6313 | 28.25 |
| 2018 | | 6331 | 28.72 |
| 2019 | | 6347 | 29.33 |
| 2020 | | 5979 | 28.09 |
| 2021 | | 2930 | 27.02 |

Table 3. Trends in smoking (years: 2011-2021; data source: NIMATS).

| ***Year of booking*** | ***Indicator level*** | ***N*** | ***%*** |
|---|---|---|---|
| 2011-2012 | Yes | 8239 | 16.74 |
| 2013-2014 | | 7241 | 15.08 |
| 2015-2016 | | 6512 | 13.73 |

| | | | |
|---|---|---|---|
| 2017-2018 | | 5992 | 13.37 |
| 2019-2021 | | 5218 | 9.64 |
| 2011-2012 | No | 40982 | 83.26 |
| 2013-2014 | | 40783 | 84.92 |
| 2015-2016 | | 40909 | 86.27 |
| 2017-2018 | | 38840 | 86.63 |
| 2019-2021 | | 48911 | 90.36 |

Table 4. Trends in periconception folic acid supplement use (years: 2015-2020; data source: NIMATS).

| *Year of booking* | *Indicator level* | *N* | *%* |
|---|---|---|---|
| 2015 | Preconception 400µg | 7975 | 34.4 |
| 2016 | | 7910 | 34.91 |
| 2017 | | 7386 | 33.43 |
| 2018 | | 7346 | 33.68 |
| 2019 | | 6922 | 32.35 |
| 2020 | | 6325 | 30.03 |
| 2015 | Preconception 5mg | 827 | 3.57 |
| 2016 | | 800 | 3.53 |
| 2017 | | 1009 | 4.57 |
| 2018 | | 1025 | 4.70 |
| 2019 | | 1192 | 5.57 |
| 2020 | | 1048 | 4.98 |
| 2015 | Postconception 400µg | 12588 | 54.31 |
| 2016 | | 11958 | 52.78 |
| 2017 | | 11603 | 52.52 |
| 2018 | | 11260 | 51.62 |
| 2019 | | 10995 | 51.38 |
| 2020 | | 11363 | 53.94 |
| 2015 | Postconception 5mg | 931 | 4.02 |
| 2016 | | 1220 | 5.38 |
| 2017 | | 1392 | 6.30 |
| 2018 | | 1435 | 6.58 |
| 2019 | | 1592 | 7.44 |
| 2020 | | 1616 | 7.67 |
| 2015 | None | 859 | 3.71 |
| 2016 | | 768 | 3.39 |
| 2017 | | 702 | 3.18 |
| 2018 | | 746 | 3.42 |
| 2019 | | 699 | 3.27 |
| 2020 | | 713 | 3.38 |

Table 5. Trends in BMI (years: 2011-2021; data source: NIMATS).

| *Year of booking* | *Indicator level* | *N* | *%* |
|---|---|---|---|
| 2011 | Underweight | 490 | 2.03 |

| | | | |
|---|---|---|---|
| 2012 | | 532 | 2.24 |
| 2013 | | 487 | 2.06 |
| 2014 | | 470 | 1.99 |
| 2015 | | 505 | 2.14 |
| 2016 | | 432 | 1.88 |
| 2017 | | 441 | 1.97 |
| 2018 | | 370 | 1.68 |
| 2019 | | 384 | 1.78 |
| 2020 | | 342 | 1.61 |
| 2021 | | 163 | 1.51 |
| 2011 | Healthy weight | 12148 | 50.27 |
| 2012 | | 11742 | 49.52 |
| 2013 | | 11656 | 49.25 |
| 2014 | | 11511 | 48.85 |
| 2015 | | 11202 | 47.39 |
| 2016 | | 10597 | 46.04 |
| 2017 | | 10213 | 45.68 |
| 2018 | | 9865 | 44.80 |
| 2019 | | 9231 | 42.73 |
| 2020 | | 8856 | 41.69 |
| 2021 | | 4316 | 39.86 |
| 2011 | Overweight | 6999 | 28.96 |
| 2012 | | 6906 | 29.12 |
| 2013 | | 6958 | 29.40 |
| 2014 | | 6828 | 28.98 |
| 2015 | | 7104 | 30.05 |
| 2016 | | 6881 | 29.89 |
| 2017 | | 6671 | 29.84 |
| 2018 | | 6541 | 29.70 |
| 2019 | | 6622 | 30.65 |
| 2020 | | 6554 | 30.85 |
| 2021 | | 3361 | 31.04 |
| 2011 | With obesity class I | 2909 | 12.04 |
| 2012 | | 2874 | 12.12 |
| 2013 | | 2885 | 12.19 |
| 2014 | | 2941 | 12.48 |
| 2015 | | 2991 | 12.65 |
| 2016 | | 3113 | 13.52 |
| 2017 | | 3071 | 13.74 |
| 2018 | | 3188 | 14.48 |
| 2019 | | 3291 | 15.23 |
| 2020 | | 3185 | 14.99 |
| 2021 | | 1747 | 16.13 |
| 2011 | With obesity class II | 1113 | 4.61 |
| 2012 | | 1112 | 4.69 |
| 2013 | | 1144 | 4.83 |

| | | | |
|---|---|---|---|
| 2014 | | 1230 | 5.22 |
| 2015 | | 1246 | 5.27 |
| 2016 | | 1341 | 5.83 |
| 2017 | | 1362 | 6.09 |
| 2018 | | 1395 | 6.33 |
| 2019 | | 1379 | 6.38 |
| 2020 | | 1547 | 7.28 |
| 2021 | | 798 | 7.37 |
| 2011 | With obesity class III | 505 | 2.09 |
| 2012 | | 548 | 2.31 |
| 2013 | | 537 | 2.27 |
| 2014 | | 583 | 2.47 |
| 2015 | | 589 | 2.49 |
| 2016 | | 655 | 2.85 |
| 2017 | | 599 | 2.68 |
| 2018 | | 662 | 3.01 |
| 2019 | | 696 | 3.22 |
| 2020 | | 761 | 3.58 |
| 2021 | | 443 | 4.09 |

### Supplementary material V

Table 1. Trends of periconception folic acid supplement use based on deprivation quintile (years: 2015-2020; data source: NIMATS).

| *Year of booking* | *Folic acid supplement use* | *Indicator level* | *N* | *%* |
|---|---|---|---|---|
| 2015 | Preconception 400µg | 1: most deprived | 1235 | 24.32 |
| 2016 | | | 1072 | 21.96 |
| 2017 | | | 988 | 20.73 |
| 2018 | | | 1027 | 21.71 |
| 2019 | | | 937 | 20.50 |
| 2020 | | | 890 | 19.45 |
| 2015 | Preconception 5mg | | 142 | 2.80 |
| 2016 | | | 156 | 3.20 |
| 2017 | | | 181 | 3.80 |
| 2018 | | | 177 | 3.74 |
| 2019 | | | 220 | 4.81 |
| 2020 | | | 177 | 3.87 |
| 2015 | Postconception 400µg | | 3144 | 61.91 |
| 2016 | | | 2994 | 61.34 |
| 2017 | | | 2976 | 62.44 |
| 2018 | | | 2867 | 60.60 |
| 2019 | | | 2766 | 60.51 |
| 2020 | | | 2833 | 61.90 |
| 2015 | Postconception 5mg | | 241 | 4.75 |
| 2016 | | | 375 | 7.68 |
| 2017 | | | 367 | 7.70 |
| 2018 | | | 373 | 7.88 |
| 2019 | | | 372 | 8.14 |
| 2020 | | | 398 | 8.70 |
| 2015 | None | | 316 | 6.22 |
| 2016 | | | 284 | 5.82 |
| 2017 | | | 254 | 5.33 |
| 2018 | | | 287 | 6.07 |
| 2019 | | | 276 | 6.04 |
| 2020 | | | 279 | 6.10 |
| 2015 | Preconception 400µg | 2 | 1609 | 32.39 |
| 2016 | | | 1556 | 31.85 |
| 2017 | | | 1467 | 31.07 |
| 2018 | | | 1426 | 30.19 |
| 2019 | | | 1330 | 29.52 |
| 2020 | | | 1232 | 27.54 |
| 2015 | Preconception 5mg | | 166 | 3.34 |
| 2016 | | | 177 | 3.62 |
| 2017 | | | 194 | 4.11 |
| 2018 | | | 205 | 4.34 |
| 2019 | | | 246 | 5.46 |
| 2020 | | | 215 | 4.81 |

| | | | | |
|---|---|---|---|---|
| 2015 | Postconception 400µg | | 2796 | 56.28 |
| 2016 | | | 2717 | 55.61 |
| 2017 | | | 2567 | 54.37 |
| 2018 | | | 2595 | 54.93 |
| 2019 | | | 2430 | 53.94 |
| 2020 | | | 2533 | 56.63 |
| 2015 | Postconception 5mg | | 200 | 4.03 |
| 2016 | | | 254 | 5.20 |
| 2017 | | | 328 | 6.95 |
| 2018 | | | 326 | 6.90 |
| 2019 | | | 344 | 7.64 |
| 2020 | | | 350 | 7.82 |
| 2015 | None | | 197 | 3.97 |
| 2016 | | | 182 | 3.72 |
| 2017 | | | 165 | 3.50 |
| 2018 | | | 172 | 3.64 |
| 2019 | | | 155 | 3.44 |
| 2020 | | | 143 | 3.20 |
| 2015 | Preconception 400µg | 3 | 1674 | 34.93 |
| 2016 | | | 1727 | 37.13 |
| 2017 | | | 1583 | 34.84 |
| 2018 | | | 1591 | 35.28 |
| 2019 | | | 1477 | 33.38 |
| 2020 | | | 1306 | 30.47 |
| 2015 | Preconception 5mg | | 171 | 3.57 |
| 2016 | | | 137 | 2.95 |
| 2017 | | | 227 | 5.00 |
| 2018 | | | 212 | 4.70 |
| 2019 | | | 231 | 5.22 |
| 2020 | | | 245 | 5.72 |
| 2015 | Postconception 400µg | | 2624 | 54.75 |
| 2016 | | | 2430 | 52.25 |
| 2017 | | | 2333 | 51.35 |
| 2018 | | | 2270 | 50.33 |
| 2019 | | | 2244 | 50.71 |
| 2020 | | | 2272 | 53.01 |
| 2015 | Postconception 5mg | | 182 | 3.80 |
| 2016 | | | 231 | 4.97 |
| 2017 | | | 273 | 6.01 |
| 2018 | | | 293 | 6.50 |
| 2019 | | | 346 | 7.82 |
| 2020 | | | 338 | 7.89 |
| 2015 | None | | 142 | 2.96 |
| 2016 | | | 126 | 2.71 |
| 2017 | | | 127 | 2.80 |
| 2018 | | | 144 | 3.19 |

| | | | | |
|---|---|---|---|---|
| 2019 | | | 127 | 2.87 |
| 2020 | | | 125 | 2.92 |
| 2015 | Preconception 400µg | 4 | 1830 | 39.48 |
| 2016 | | | 1821 | 39.95 |
| 2017 | | | 1630 | 37.83 |
| 2018 | | | 1685 | 39.15 |
| 2019 | | | 1575 | 37.29 |
| 2020 | | | 1444 | 34.78 |
| 2015 | Preconception 5mg | | 193 | 4.16 |
| 2016 | | | 177 | 3.88 |
| 2017 | | | 226 | 5.24 |
| 2018 | | | 225 | 5.23 |
| 2019 | | | 263 | 6.23 |
| 2020 | | | 215 | 5.18 |
| 2015 | Postconception 400µg | | 2302 | 49.67 |
| 2016 | | | 2241 | 49.17 |
| 2017 | | | 2107 | 48.90 |
| 2018 | | | 2045 | 47.51 |
| 2019 | | | 1978 | 46.83 |
| 2020 | | | 2079 | 50.07 |
| 2015 | Postconception 5mg | | 176 | 3.80 |
| 2016 | | | 209 | 4.59 |
| 2017 | | | 240 | 5.57 |
| 2018 | | | 259 | 6.02 |
| 2019 | | | 311 | 7.36 |
| 2020 | | | 313 | 7.54 |
| 2015 | None | | 134 | 2.89 |
| 2016 | | | 110 | 2.41 |
| 2017 | | | 106 | 2.46 |
| 2018 | | | 90 | 2.09 |
| 2019 | | | 97 | 2.30 |
| 2020 | | | 101 | 2.43 |
| 2015 | Preconception 400µg | 5: least deprived | 1627 | 43.9 |
| 2016 | | | 1734 | 47.12 |
| 2017 | | | 1718 | 45.78 |
| 2018 | | | 1617 | 45.64 |
| 2019 | | | 1603 | 43.62 |
| 2020 | | | 1453 | 40.62 |
| 2015 | Preconception 5mg | | 155 | 4.18 |
| 2016 | | | 153 | 4.16 |
| 2017 | | | 181 | 4.82 |
| 2018 | | | 206 | 5.81 |
| 2019 | | | 232 | 6.31 |
| 2020 | | | 196 | 5.48 |
| 2015 | Postconception 400µg | | 1722 | 46.47 |
| 2016 | | | 1576 | 42.83 |

| | | | | |
|---|---|---|---|---|
| 2017 | | | 1620 | 43.17 |
| 2018 | | | 1483 | 41.86 |
| 2019 | | | 1577 | 42.91 |
| 2020 | | | 1646 | 46.02 |
| 2015 | Postconception 5mg | | 132 | 3.56 |
| 2016 | | | 151 | 4.10 |
| 2017 | | | 184 | 4.90 |
| 2018 | | | 184 | 5.19 |
| 2019 | | | 219 | 5.96 |
| 2020 | | | 217 | 6.07 |
| 2015 | None | | 70 | 1.89 |
| 2016 | | | 66 | 1.79 |
| 2017 | | | 50 | 1.33 |
| 2018 | | | 53 | 1.50 |
| 2019 | | | 44 | 1.20 |
| 2020 | | | 65 | 1.82 |

Table 2. Trends of periconception folic acid supplement use based on age at booking (years: 2015-2020; data source: NIMATS).

| ***Year of booking*** | ***Folic acid supplement use*** | ***Indicator level*** | ***N*** | ***%*** |
|---|---|---|---|---|
| 2015 | Preconception 400µg | <25Y | 567 | 12.87 |
| 2016 | | | 526 | 13.14 |
| 2017 | | | 437 | 11.36 |
| 2018 | | | 418 | 11.16 |
| 2019 | | | 411 | 11.67 |
| 2020 | | | 383 | 11.80 |
| 2015 | Preconception 5mg | | 64 | 1.45 |
| 2016 | | | 69 | 1.72 |
| 2017 | | | 78 | 2.03 |
| 2018 | | | 87 | 2.32 |
| 2019 | | | 86 | 2.44 |
| 2020 | | | 77 | 2.37 |
| 2015 | Postconception 400µg | | 3232 | 73.39 |
| 2016 | | | 2883 | 72.02 |
| 2017 | | | 2831 | 73.59 |
| 2018 | | | 2679 | 71.54 |
| 2019 | | | 2502 | 71.06 |
| 2020 | | | 2320 | 71.49 |
| 2015 | Postconception 5mg | | 200 | 4.54 |
| 2016 | | | 256 | 6.40 |
| 2017 | | | 257 | 6.68 |
| 2018 | | | 276 | 7.37 |
| 2019 | | | 273 | 7.75 |
| 2020 | | | 226 | 6.96 |
| 2015 | None | | 341 | 7.74 |

| | | | | |
|---|---|---|---|---|
| 2016 | | | 269 | 6.72 |
| 2017 | | | 244 | 6.34 |
| 2018 | | | 285 | 7.61 |
| 2019 | | | 249 | 7.07 |
| 2020 | | | 239 | 7.37 |
| 2015 | Preconception 400µg | 25-34Y | 5474 | 38.07 |
| 2016 | | | 5479 | 38.44 |
| 2017 | | | 5087 | 36.84 |
| 2018 | | | 5075 | 37.27 |
| 2019 | | | 4749 | 35.78 |
| 2020 | | | 4283 | 32.07 |
| 2015 | Preconception 5mg | | 524 | 3.64 |
| 2016 | | | 484 | 3.40 |
| 2017 | | | 632 | 4.58 |
| 2018 | | | 602 | 4.42 |
| 2019 | | | 692 | 5.21 |
| 2020 | | | 656 | 4.91 |
| 2015 | Postconception 400µg | | 7418 | 51.59 |
| 2016 | | | 7188 | 50.44 |
| 2017 | | | 6902 | 49.99 |
| 2018 | | | 6723 | 49.38 |
| 2019 | | | 6546 | 49.32 |
| 2020 | | | 7056 | 52.83 |
| 2015 | Postconception 5mg | | 561 | 3.90 |
| 2016 | | | 718 | 5.04 |
| 2017 | | | 820 | 5.94 |
| 2018 | | | 848 | 6.23 |
| 2019 | | | 938 | 7.07 |
| 2020 | | | 992 | 7.43 |
| 2015 | None | | 401 | 2.79 |
| 2016 | | | 383 | 2.69 |
| 2017 | | | 367 | 2.66 |
| 2018 | | | 368 | 2.70 |
| 2019 | | | 347 | 2.61 |
| 2020 | | | 368 | 2.76 |
| 2015 | Preconception 400µg | 35+Y | 1934 | 43.97 |
| 2016 | | | 1905 | 43.29 |
| 2017 | | | 1862 | 41.97 |
| 2018 | | | 1853 | 41.63 |
| 2019 | | | 1762 | 38.25 |
| 2020 | | | 1659 | 37.16 |
| 2015 | Preconception 5mg | | 239 | 5.43 |
| 2016 | | | 247 | 5.61 |
| 2017 | | | 299 | 6.74 |
| 2018 | | | 336 | 7.55 |
| 2019 | | | 414 | 8.99 |

| | | | | |
|---|---|---|---|---|
| 2020 | | | 315 | 7.05 |
| 2015 | Postconception 400µg | | 1938 | 44.07 |
| 2016 | | | 1887 | 42.88 |
| 2017 | | | 1870 | 42.15 |
| 2018 | | | 1858 | 41.74 |
| 2019 | | | 1947 | 42.26 |
| 2020 | | | 1987 | 44.5 |
| 2015 | Postconception 5mg | | 170 | 3.87 |
| 2016 | | | 246 | 5.59 |
| 2017 | | | 315 | 7.10 |
| 2018 | | | 311 | 6.99 |
| 2019 | | | 381 | 8.27 |
| 2020 | | | 398 | 8.91 |
| 2015 | None | | 117 | 2.66 |
| 2016 | | | 116 | 2.64 |
| 2017 | | | 91 | 2.05 |
| 2018 | | | 93 | 2.09 |
| 2019 | | | 103 | 2.24 |
| 2020 | | | 106 | 2.37 |

Table 3. Trends of periconception folic acid supplement use based on maternal gravida (years: 2015-2020; data source: NIMATS).

| *Year of booking* | *Folic acid supplement use* | *Indicator level* | *N* | *%* |
|---|---|---|---|---|
| 2015 | Preconception 400µg | 1: first-time pregnancy | 2473 | 34.52 |
| 2016 | | | 2605 | 37.48 |
| 2017 | | | 2424 | 35.59 |
| 2018 | | | 2373 | 35.03 |
| 2019 | | | 2378 | 34.74 |
| 2020 | | | 2071 | 32.71 |
| 2015 | Preconception 5mg | | 194 | 2.71 |
| 2016 | | | 221 | 3.18 |
| 2017 | | | 231 | 3.39 |
| 2018 | | | 272 | 4.01 |
| 2019 | | | 303 | 4.43 |
| 2020 | | | 258 | 4.08 |
| 2015 | Postconception 400µg | | 3974 | 55.48 |
| 2016 | | | 3605 | 51.87 |
| 2017 | | | 3651 | 53.60 |
| 2018 | | | 3542 | 52.28 |
| 2019 | | | 3541 | 51.73 |
| 2020 | | | 3415 | 53.94 |
| 2015 | Postconception 5mg | | 269 | 3.76 |
| 2016 | | | 333 | 4.79 |
| 2017 | | | 361 | 5.30 |
| 2018 | | | 384 | 5.67 |

| | | | | |
|---|---|---|---|---|
| 2019 | | | 443 | 6.47 |
| 2020 | | | 389 | 6.14 |
| 2015 | None | | 253 | 3.53 |
| 2016 | | | 186 | 2.68 |
| 2017 | | | 144 | 2.11 |
| 2018 | | | 204 | 3.01 |
| 2019 | | | 180 | 2.63 |
| 2020 | | | 198 | 3.13 |
| 2015 | Preconception 400µg | ≥2: subsequent pregnancy | 5502 | 34.35 |
| 2016 | | | 5305 | 33.78 |
| 2017 | | | 4962 | 32.47 |
| 2018 | | | 4973 | 33.07 |
| 2019 | | | 4544 | 31.22 |
| 2020 | | | 4254 | 28.87 |
| 2015 | Preconception 5mg | | 633 | 3.95 |
| 2016 | | | 579 | 3.69 |
| 2017 | | | 778 | 5.09 |
| 2018 | | | 753 | 5.01 |
| 2019 | | | 889 | 6.11 |
| 2020 | | | 790 | 5.36 |
| 2015 | Postconception 400µg | | 8614 | 53.78 |
| 2016 | | | 8353 | 53.18 |
| 2017 | | | 7952 | 52.04 |
| 2018 | | | 7718 | 51.33 |
| 2019 | | | 7454 | 51.21 |
| 2020 | | | 7948 | 53.94 |
| 2015 | Postconception 5mg | | 662 | 4.13 |
| 2016 | | | 887 | 5.65 |
| 2017 | | | 1031 | 6.75 |
| 2018 | | | 1051 | 6.99 |
| 2019 | | | 1149 | 7.89 |
| 2020 | | | 1227 | 8.33 |
| 2015 | None | | 606 | 3.78 |
| 2016 | | | 582 | 3.71 |
| 2017 | | | 558 | 3.65 |
| 2018 | | | 542 | 3.60 |
| 2019 | | | 519 | 3.57 |
| 2020 | | | 515 | 3.50 |

Table 4. Trends of periconception folic acid supplement use based on planned pregnancy (years: 2015-2020; data source: NIMATS).

| ***Year of booking*** | ***Folic acid supplement use*** | ***Indicator level*** | ***N*** | ***%*** |
|---|---|---|---|---|
| 2015 | Preconception 400µg | Yes | 7596 | 45.35 |
| 2016 | | | 7541 | 46.00 |
| 2017 | | | 6993 | 44.15 |

| | | | | |
|---|---|---|---|---|
| 2018 | | | 7010 | 45.10 |
| 2019 | | | 6525 | 43.16 |
| 2020 | | | 5990 | 39.54 |
| 2015 | Preconception 5mg | | 753 | 4.50 |
| 2016 | | | 728 | 4.44 |
| 2017 | | | 897 | 5.66 |
| 2018 | | | 914 | 5.88 |
| 2019 | | | 1063 | 7.03 |
| 2020 | | | 936 | 6.18 |
| 2015 | Postconception 400µg | | 7498 | 44.76 |
| 2016 | | | 7088 | 43.24 |
| 2017 | | | 6842 | 43.20 |
| 2018 | | | 6485 | 41.73 |
| 2019 | | | 6316 | 41.78 |
| 2020 | | | 6899 | 45.53 |
| 2015 | Postconception 5mg | | 546 | 3.26 |
| 2016 | | | 722 | 4.40 |
| 2017 | | | 842 | 5.32 |
| 2018 | | | 825 | 5.31 |
| 2019 | | | 973 | 6.44 |
| 2020 | | | 1015 | 6.70 |
| 2015 | None | | 357 | 2.13 |
| 2016 | | | 314 | 1.92 |
| 2017 | | | 264 | 1.67 |
| 2018 | | | 308 | 1.98 |
| 2019 | | | 241 | 1.59 |
| 2020 | | | 311 | 2.05 |
| 2015 | Preconception 400µg | No | 379 | 5.89 |
| 2016 | | | 369 | 5.89 |
| 2017 | | | 393 | 6.28 |
| 2018 | | | 336 | 5.36 |
| 2019 | | | 397 | 6.32 |
| 2020 | | | 335 | 5.66 |
| 2015 | Preconception 5mg | | 74 | 1.15 |
| 2016 | | | 72 | 1.15 |
| 2017 | | | 112 | 1.79 |
| 2018 | | | 111 | 1.77 |
| 2019 | | | 129 | 2.05 |
| 2020 | | | 112 | 1.89 |
| 2015 | Postconception 400µg | | 5090 | 79.16 |
| 2016 | | | 4870 | 77.76 |
| 2017 | | | 4761 | 76.13 |
| 2018 | | | 4775 | 76.16 |
| 2019 | | | 4679 | 74.48 |
| 2020 | | | 4464 | 75.48 |
| 2015 | Postconception 5mg | | 385 | 5.99 |

| | | | | |
|---|---|---|---|---|
| 2016 | | | 498 | 7.95 |
| 2017 | | | 550 | 8.79 |
| 2018 | | | 610 | 9.73 |
| 2019 | | | 619 | 9.85 |
| 2020 | | | 601 | 10.16 |
| 2015 | None | | 502 | 7.81 |
| 2016 | | | 454 | 7.25 |
| 2017 | | | 438 | 7.00 |
| 2018 | | | 438 | 6.99 |
| 2019 | | | 458 | 7.29 |
| 2020 | | | 402 | 6.80 |

Table 5. Trends of periconception folic acid supplement use based on maternal BMI category (years: 2015-2020; data source: NIMATS).

| *Year of booking* | *Folic acid supplement use* | *Indicator level* | *N* | *%* |
|---|---|---|---|---|
| 2015-2016 | Preconception 400µg | Underweight | 257 | 28.00 |
| 2017-2018 | | | 208 | 26.00 |
| 2019-2020 | | | 179 | 24.9 |
| 2015-2016 | Preconception 5mg | | 21 | 2.29 |
| 2017-2018 | | | 14 | 1.75 |
| 2019-2020 | | | 16 | 2.23 |
| 2015-2016 | Postconception 400µg | | 574 | 62.53 |
| 2017-2018 | | | 506 | 63.25 |
| 2019-2020 | | | 463 | 64.39 |
| 2015-2016 | Postconception 5mg | | 22 | 2.4 |
| 2017-2018 | | | 32 | 4.00 |
| 2019-2020 | | | 27 | 3.76 |
| 2015-2016 | None | | 44 | 4.79 |
| 2017-2018 | | | 40 | 5.00 |
| 2019-2020 | | | 34 | 4.73 |
| 2015-2016 | Preconception 400µg | Healthy weight | 8012 | 37.46 |
| 2017-2018 | | | 7519 | 37.85 |
| 2019-2020 | | | 6473 | 36.14 |
| 2015-2016 | Preconception 5mg | | 597 | 2.79 |
| 2017-2018 | | | 621 | 3.13 |
| 2019-2020 | | | 551 | 3.08 |
| 2015-2016 | Postconception 400µg | | 11566 | 54.07 |
| 2017-2018 | | | 10528 | 53.00 |
| 2019-2020 | | | 9828 | 54.87 |
| 2015-2016 | Postconception 5mg | | 530 | 2.48 |
| 2017-2018 | | | 567 | 2.85 |
| 2019-2020 | | | 510 | 2.85 |
| 2015-2016 | None | | 685 | 3.20 |
| 2017-2018 | | | 630 | 3.17 |
| 2019-2020 | | | 550 | 3.07 |

| | | | | |
|---|---|---|---|---|
| 2015-2016 | Preconception 400µg | Overweight | 4904 | 35.71 |
| 2017-2018 | | | 4563 | 34.94 |
| 2019-2020 | | | 4366 | 33.40 |
| 2015-2016 | Preconception 5mg | | 410 | 2.99 |
| 2017-2018 | | | 496 | 3.80 |
| 2019-2020 | | | 523 | 4.00 |
| 2015-2016 | Postconception 400µg | | 7507 | 54.66 |
| 2017-2018 | | | 7086 | 54.26 |
| 2019-2020 | | | 7266 | 55.59 |
| 2015-2016 | Postconception 5mg | | 408 | 2.97 |
| 2017-2018 | | | 473 | 3.62 |
| 2019-2020 | | | 480 | 3.67 |
| 2015-2016 | None | | 505 | 3.68 |
| 2017-2018 | | | 442 | 3.38 |
| 2019-2020 | | | 436 | 3.34 |
| 2015-2016 | Preconception 400µg | With obesity class I | 1825 | 30.34 |
| 2017-2018 | | | 1705 | 27.53 |
| 2019-2020 | | | 1588 | 24.74 |
| 2015-2016 | Preconception 5mg | | 318 | 5.29 |
| 2017-2018 | | | 447 | 7.22 |
| 2019-2020 | | | 549 | 8.55 |
| 2015-2016 | Postconception 400µg | | 3100 | 51.54 |
| 2017-2018 | | | 3107 | 50.16 |
| 2019-2020 | | | 3177 | 49.5 |
| 2015-2016 | Postconception 5mg | | 517 | 8.60 |
| 2017-2018 | | | 731 | 11.8 |
| 2019-2020 | | | 874 | 13.62 |
| 2015-2016 | None | | 255 | 4.24 |
| 2017-2018 | | | 204 | 3.29 |
| 2019-2020 | | | 230 | 3.58 |
| 2015-2016 | Preconception 400µg | With obesity class II | 641 | 25.13 |
| 2017-2018 | | | 555 | 20.33 |
| 2019-2020 | | | 483 | 16.66 |
| 2015-2016 | Preconception 5mg | | 179 | 7.02 |
| 2017-2018 | | | 290 | 10.62 |
| 2019-2020 | | | 374 | 12.90 |
| 2015-2016 | Postconception 400µg | | 1244 | 48.77 |
| 2017-2018 | | | 1168 | 42.78 |
| 2019-2020 | | | 1168 | 40.29 |
| 2015-2016 | Postconception 5mg | | 391 | 15.33 |
| 2017-2018 | | | 622 | 22.78 |
| 2019-2020 | | | 768 | 26.49 |
| 2015-2016 | None | | 96 | 3.76 |
| 2017-2018 | | | 95 | 3.48 |
| 2019-2020 | | | 106 | 3.66 |
| 2015-2016 | Preconception 400µg | With obesity class III | 246 | 20.03 |

| | | | | |
|---|---|---|---|---|
| 2017-2018 | | | 182 | 14.50 |
| 2019-2020 | | | 158 | 10.93 |
| 2015-2016 | Preconception 5mg | | 102 | 8.31 |
| 2017-2018 | | | 166 | 13.23 |
| 2019-2020 | | | 227 | 15.70 |
| 2015-2016 | Postconception 400µg | | 555 | 45.20 |
| 2017-2018 | | | 468 | 37.29 |
| 2019-2020 | | | 456 | 31.54 |
| 2015-2016 | Postconception 5mg | | 283 | 23.05 |
| 2017-2018 | | | 402 | 32.03 |
| 2019-2020 | | | 549 | 37.97 |
| 2015-2016 | None | | 42 | 3.42 |
| 2017-2018 | | | 37 | 2.95 |
| 2019-2020 | | | 56 | 3.87 |

### Supplementary material VI

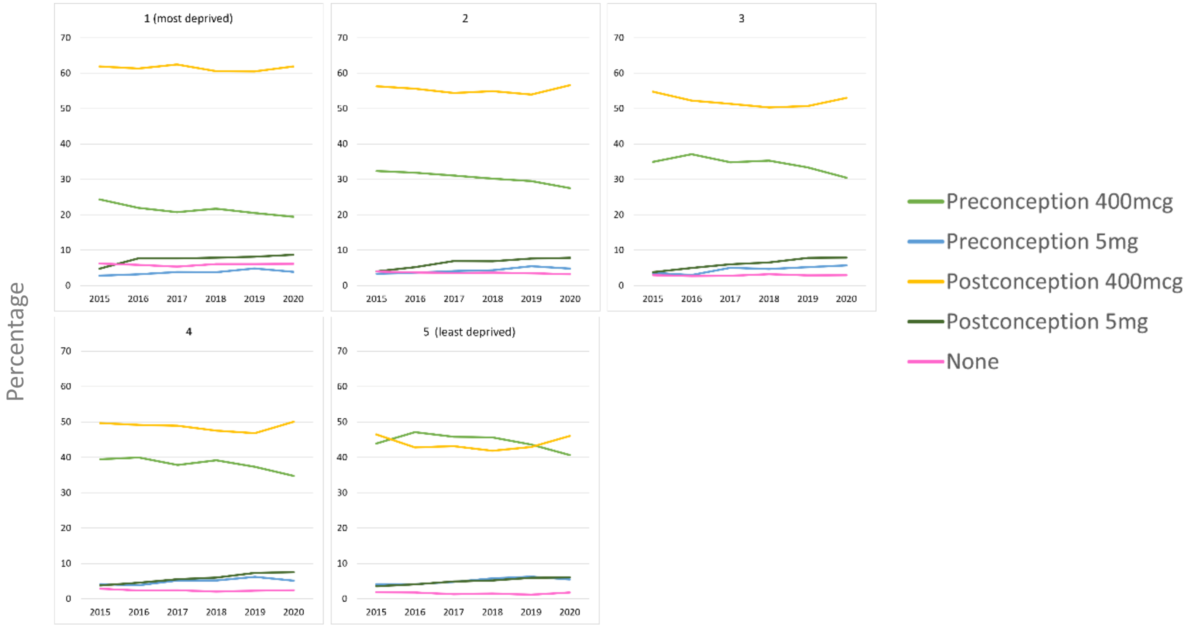


Figure 1a. Prevalence of folic acid supplement use levels based on deprivation quintile.

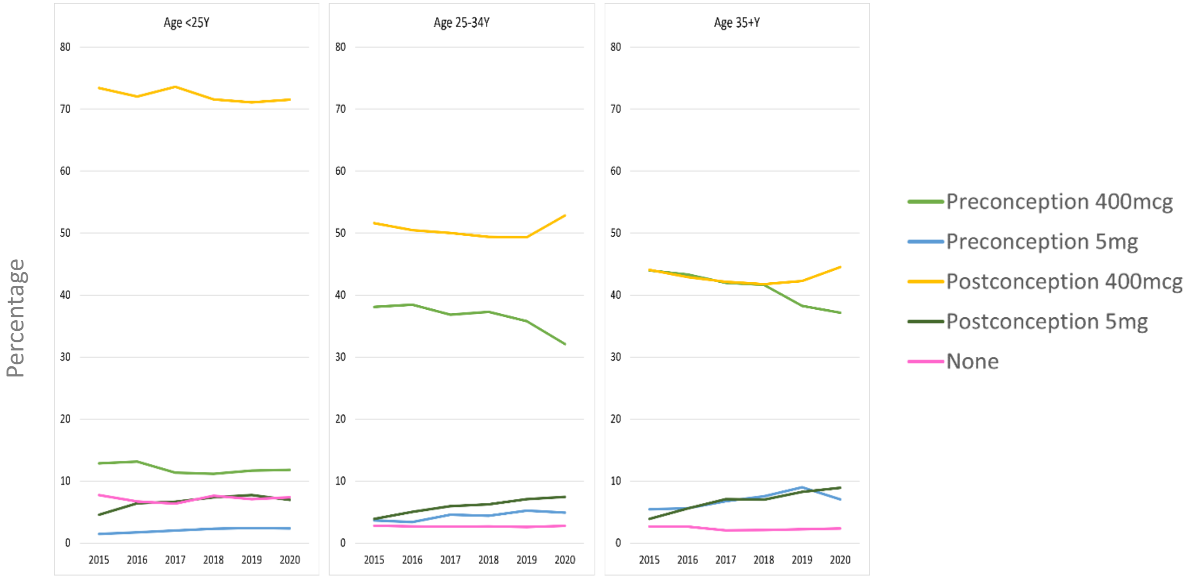


Figure 1b. Prevalence of folic acid supplement use levels based on age at booking.

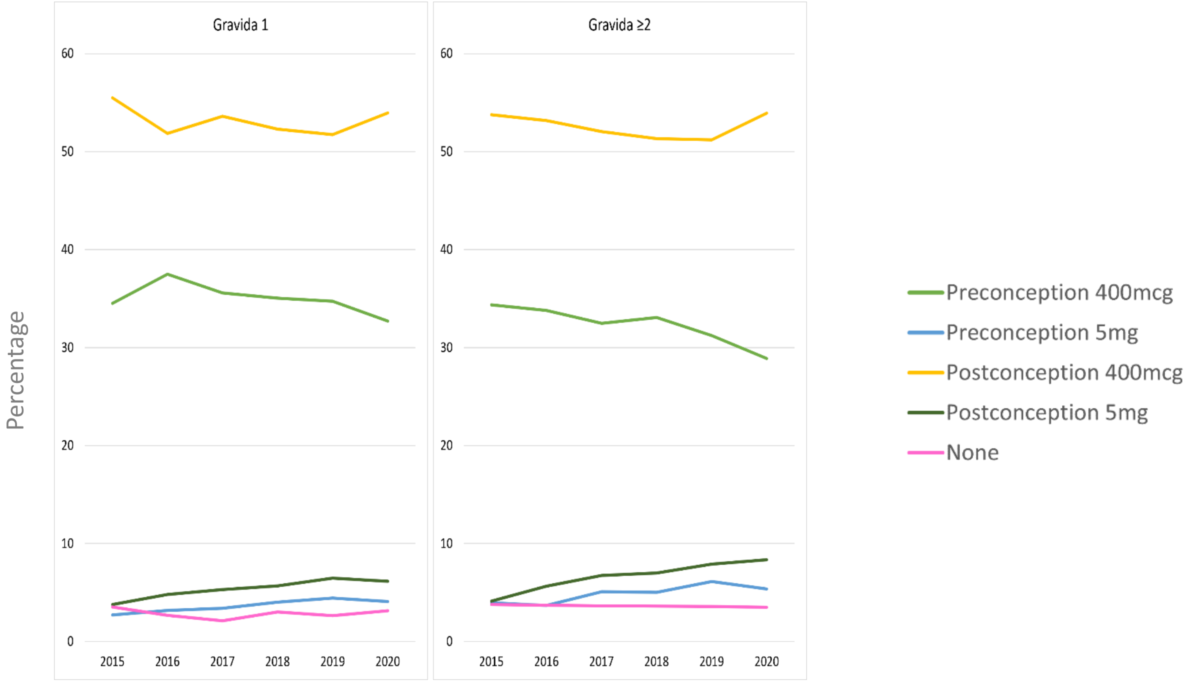


Figure 1c. Prevalence of folic acid supplement use levels based on maternal gravidity.

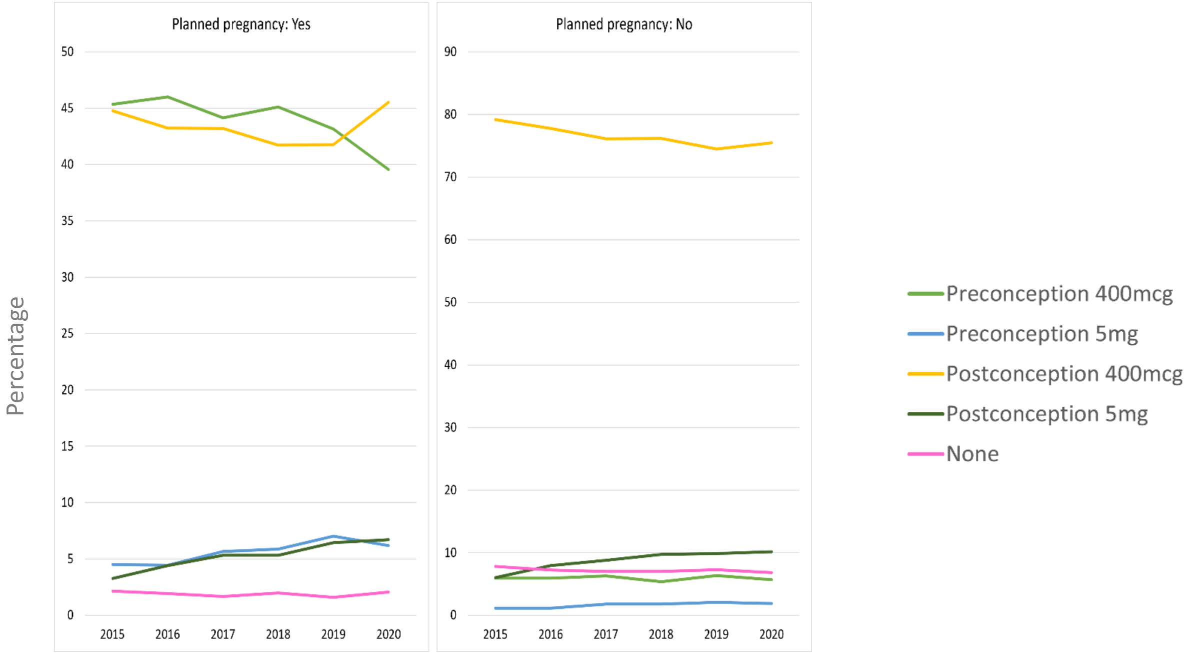


Figure 1d. Prevalence of folic acid supplement use levels based on planned pregnancies.

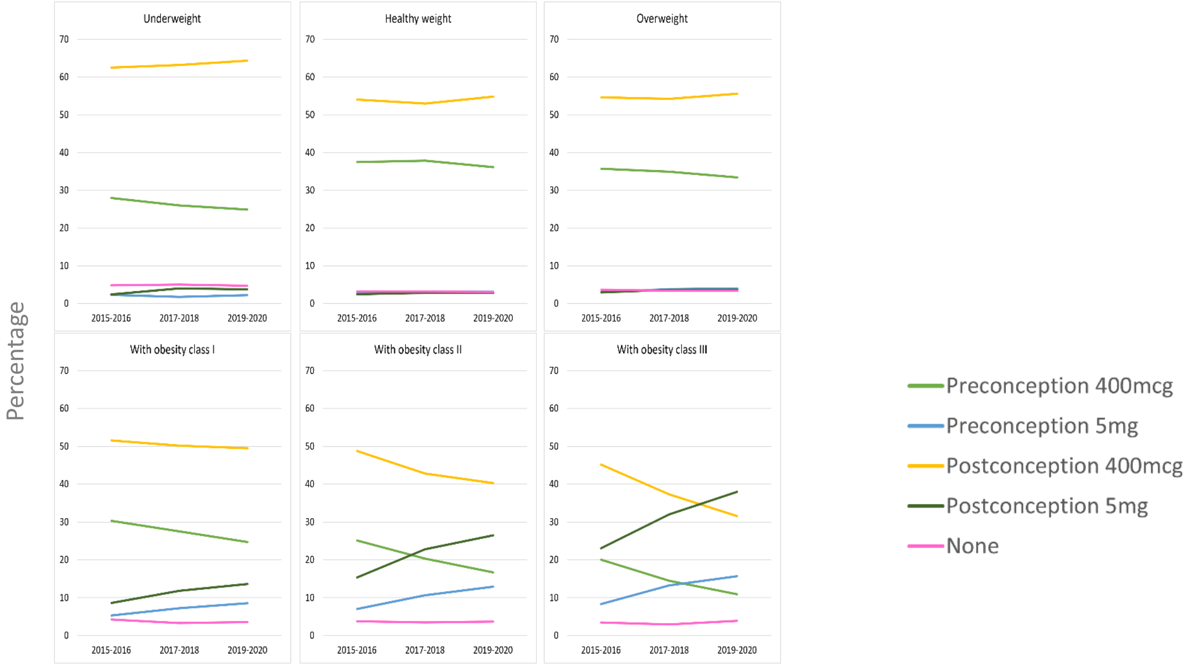


Figure 1e. Prevalence of folic acid supplement use levels based on maternal BMI.

### Supplementary material VII

Box 1. Summary of key discussions discussing folic acid supplement use in NI with PPIE contributors.

*Understanding folic acid supplement use trends: discussion on preparation for pregnancy*

Some women recalled receiving generic preconception advice pertaining to nutritional optimisation (e.g., 'avoiding sugary stuff') or 'slowing down' to avoid excessive stress and anxieties, sometimes from family members. When discussing folic acid supplement use specifically, there was some acknowledgement among the group that this is an important behaviour to engage with. However, some women were unsure what folic acid supplements are recommended for and some were worried about side effects. Most women were not aware that there are two doses available in the UK (i.e., 400µg and 5mg).

Many women acknowledged the challenge presented by unplanned pregnancies and the disinclination to disclose pregnancy intentions prior to conception.

*Sources of information*

Some women discussed looking for information online in preparation for their pregnancy (e.g., NHS website), with some recalling that family members also recommended folic acid supplement use before conception. Healthcare professionals were highlighted as key sources of information, though two women in particular expressed receiving little support before and in the early stages of pregnancy, and described their HCP as 'judgemental', particularly in relation to body weight. Other women agreed that GPs are not always easily approachable.

TikTok was mentioned as a platform for advice/information retrieval; it was described as more up-to-date than other platforms, including YouTube. However, there was also some distrust in online advice, especially from these types of social media, with a woman stating that 'old information is recycled' when it comes to preconception advice.

*Suggestions to improve folic acid supplement use*

Suggestions to improve the provision of folic acid supplement use advice initially revolved around raising awareness via GPs, including flyers and posters within GP surgeries. Women wanted more personalised care from GPs ('no one size fits all') and more educational information to support their decisions. The limited time dedicated to each patient by GPs was also acknowledged, alongside the recognition that they are under pressure within the healthcare system. The importance of providing folic acid supplement use education in school was mentioned. However, some women revealed they might not have been interested in learning about this topic at school age, feeling like it was not relevant for them at that life stage.

The role of men was not openly discussed, with women ultimately stating they felt the main responsibilities lay with them (and with that also the blame in case of adverse pregnancy outcomes). However, when prompted, some did acknowledge that men should also be involved in the optimisation of health before conception.

Supported by our findings, women recognised that some groups of women (e.g., younger women, non-planners) may need extra support in regard to preconception folic acid supplement use.

Abbreviations: GP: General Practitioner; HCP: Healthcare professional.
